# Raising the bar for patient experience during care transitions in Canada: A repeated cross-sectional survey evaluating a patient-oriented discharge summary at Ontario hospitals

**DOI:** 10.1101/2022.05.01.22274547

**Authors:** Karen Okrainec, Audrey Chaput, Valeria E. Rac, George Tomlinson, John Matelski, Mark Robson, Amy Troup, Murray Krahn, Shoshana Hahn-Goldberg

## Abstract

**Background:** Patient experience when transitioning home from hospital is an important quality metric linked to improved patient outcomes. We evaluated the impact of a hospital-based care transition intervention, patient-oriented discharge summary (PODS), on patient experience across Ontario acute care hospitals.

**Methods:** We used a repeated cross-sectional study design to compare yearly positive responses to four questions centered on discharge communication from the Canadian Patient Experience Survey (2016-2020) among three hospital cohorts with various levels of PODS implementation. Logistic regression using a binomial likelihood accounting for site level clustering was used to assess continuous linear time trends among cohorts and cohort differences during the post-implementation period. This research had oversight from a public advisory group of patient and caregiver partners from across the province.

**Results:** 512,288 responses were included with mean age 69 ± 14 years (females) and 61 ± 20 years (males). Compared to non-implementation hospitals, hospitals with full implementation (>50% discharges) reported higher odds for having discussed the help needed when leaving hospital (OR=1.18, 95% CI=1.02-1.37) and having received information in writing about what symptoms to look out for (OR=1.44, 95%=1.17-1.78) post-implementation. The linear time trend was also significant when comparing hospitals with full versus no implementation for having received information in writing about what symptoms to look out for (OR=1.05, 95% CI=1.01-1.09).

**Interpretation:** PODS implementation was associated with higher odds of positive patient experience, particularly for discharge planning. Further efforts should center on discharge management, specifically: understanding of medications and what to do if worried once home.

## Introduction

Improving patient experience following hospital admission is an important target of health systems worldwide [1-3]. Patient experience is a patient-reported experience measure that allows patients to rank various elements of the hospitalization that were important to them using standardized and validated measures that can be compared across patients, institutions, provinces, countries and drive improvements in quality of care [4]. Patient experience is often also used to complement patient-reported outcome measures to help understand rising numbers of avoidable health-care utilization, yet rarely considered when evaluating the impact of care transition interventions or new models of care [5]. The Hospital Consumer Assessment of Healthcare Providers and Systems (HCAHPS) survey, designed in the US, is becoming increasingly recognized and used in many parts of the world [6-8] along with the three-item version of the Care Transitions Measure (CTM-3) [9]. In 2014, the Canadian Institute for Health Information created the first Canadian Patient Experience - Inpatient Care (CPES-IC) survey to capture the quality of communication, information sharing and other markers of patient experience for Canadian patients discharged from acute care hospitals [10].

The Patient Oriented-Discharge Summary (PODS) is a novel individualized discharge instruction tool containing five sections of information centered on changes to medications, daily activities and diet, follow-up appointments, symptoms to watch out for and resources [11]. Co-designed with patients and caregivers to improve communication, PODS has been found to increase patient-centered discharge practices by involving family caregivers in discharge conversations and by increasing the use of teach-back, two communication practices known to improve patient outcomes [12,13]. The objective of this study was to evaluate the association of PODS with patient experience among surgical and medical inpatients across Ontario hospitals.

## Methods

Between April 2017 and March 2018, 21 hospitals in Ontario implemented PODS in an inpatient population of their choice through a funded and supported community of practice [13]. The study was reviewed by the University Health Network (UHN) research ethics board. The UHN REB waived ethics approval given the use of aggregate non patient identified data (Waiver 17-5469). Consent was not obtained as the data were analyzed anonymously.

### Intervention

PODS contains five sections of written discharge instructions co-designed with patients and family caregivers to be meaningful and usable: 1) medications, 2) changes to daily activities and diet, follow-up appointments, 4) expected and worrisome symptoms to watch for after leaving hospital and 5) resources and contact information. PODS includes design features (large font, pictograms, note-taking section) known to enhance retention and understanding of instructions and is accompanied by process guidelines which foster patient and caregiver engagement and teach-back when reviewing discharge instructions [12]. Prior to PODS implementation, discharge instructions in Ontario hospitals were verbal or a summary directed to the primary care provider [11].

### Setting/Patients

Ontario is Canada’s largest province which includes 123 acute care hospitals. The CPES-IC is a standardized and validated survey administered by email, mail or telephone in English or French to adults over 18 years discharged from an Ontario acute care hospital’s medical or surgical unit in the previous 48 hours to three months [14]. Ontario hospitals who collected patient experience data among their medical or surgical inpatients during all four years of the study period, April 2016 to March 2020, were eligible for inclusion in our study.

We received anonymized data to the CPES-IC survey for all hospitals by pre-assigned cohorts. The first cohort included hospitals who implemented PODS among ≥ 50% of medical and surgical inpatients between April 1, 2017 and March 2018. Our second cohort included hospitals who had implemented PODS among <50% of their medical or surgical inpatients in our implementation study by March 2018 or who had some known implementation outside this study. Our third cohort included all other hospitals who collected outcome data but who had no PODS implementation as of December 2018. Further data on yearly mean age, mean proportion of sex represented by the CPES-IC survey for respondents along with yearly response rate for aggregate questions were obtained from the Canadian Institute for Health Information.

### Design

Our study used a repeated cross-sectional design to measure outcomes at four different time points: 1) pre-implementation (April 1, 2016 to March 31, 2017), 2) year 1 of implementation (April 1, 2017 to March 31, 2018), 3) year 2 of implementation (April 1, 2018 to March 31, 2019) and 4) post-implementation (April 1, 2019 to March 31, 2020) in all three cohorts. We used the STROBE cross-sectional checklist when writing our report [15].

### Outcomes

We used individual responses to the following four questions from CPES-IC centered on discharge communication as they represented content directly addressed in the PODS intervention. To reflect <u>discharge planning,</u> responses to the following two questions were used: 1) During this hospital stay, did doctors, nurses or other hospital staff talk with you about whether you would have the help needed when you left hospital? Responses included no or yes; 2) During this hospital stay, did you get information in writing about what symptoms or health problems to look out for after you left hospital? Responses included no or yes. To reflect <u>discharge management</u>, responses to the following two questions were used: 3) Before you left the hospital, did you have a clear understanding about all your prescribed medications, including those you were taking before your hospital stay? Responses included not at all, partly, quite a bit, completely, not applicable; 4) Did you receive enough information from hospital staff about what to do if you were worried about your condition or treatment after you left the hospital? Responses included not at all, partly, quite a bit, and completely.

### Statistical Analysis

For each survey question, aggregate percentages of the top response and 95% confidence intervals for these estimates were computed for each cohort at each time point using Generalized Estimating Equations (GEE), which allowed us to account for site level clustering. A second GEE model treated time as a continuous predictor to evaluate linear time trends for each cohort and to assess cohort level differences in the post implementation period. We report all estimates as odds ratios, along with 95% confidence intervals and Wald test p-values. Alpha= .05 is used as the threshold for statistical significance. The models were fit using the geepack package in R version 3.6.2.

### Involvement of Patient Partners

Our study involved patients and caregivers as research partners. One of our caregiver partners (AC) has been a research partner in all aspects of the study, including as principal applicant on the grant which funded this work and identifying original research questions and outcomes for this study. Subsequently, AC recruited four other patient and caregiver partners to join a provincial advisory group in June 2020. This advisory group (AC, AF, GC, TM, and JR) subsequently collaborated in this work by reviewing results and through discussion, informed the framing of our paper, including the discussion and conclusions. AC subsequently critically reviewed the final version and is a co-author on this paper.

## Results

A total of 512,288 responses aggregated from 59 hospitals were analyzed. Cohort 1 included eight hospitals who implemented PODS fully, cohort 2 included 16 hospitals who implemented PODS partially and cohort 3 included 36 hospitals who had no PODS implementation. The mean age by gender and year for all respondents in Ontario is listed in Table 1. Response rates across Ontario hospitals to the Patient Experience Survey ranged between 28% and 35% (Table 2).

**Table 1.**
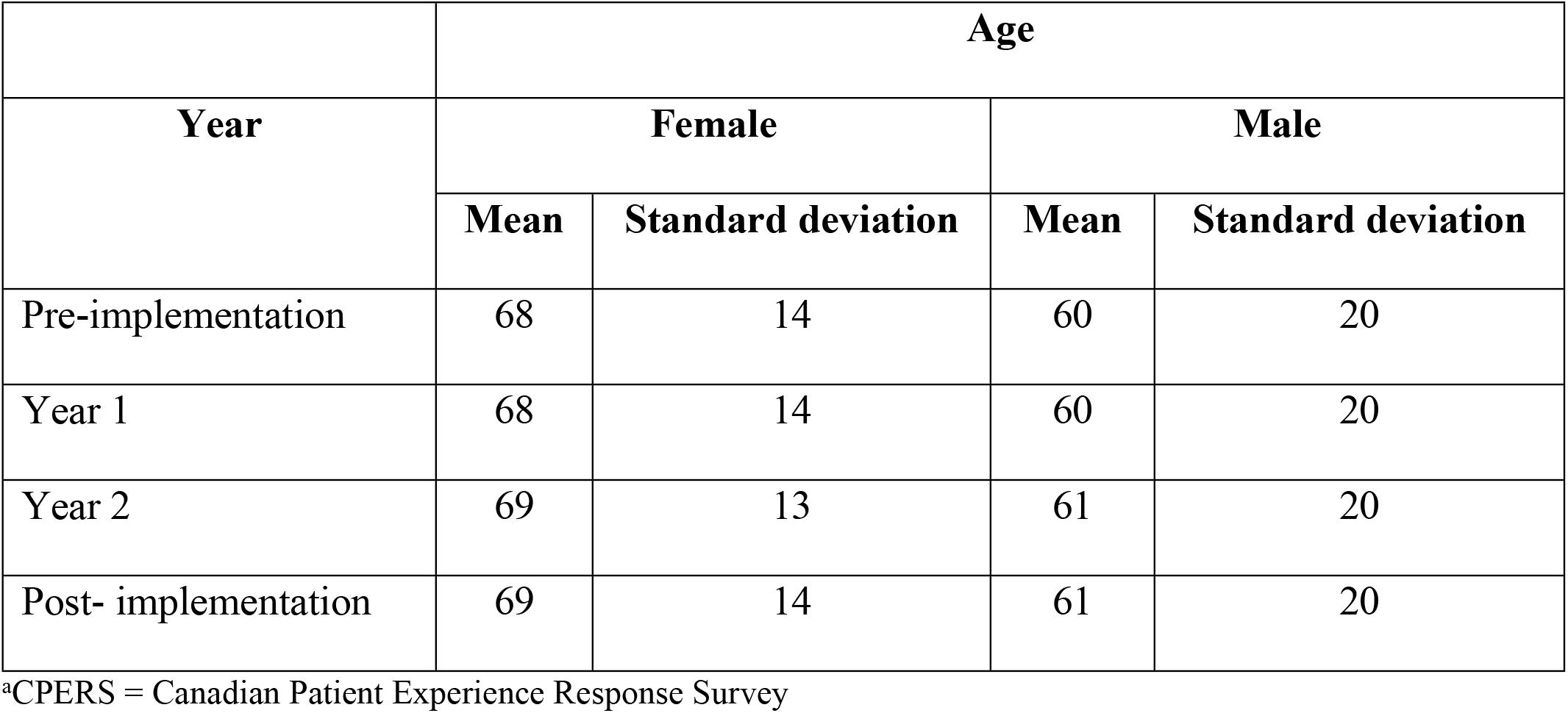
Mean Age of Ontario Patients in CPERS^a^ by Gender and Fiscal Year

| Year | Age |  |  |  |
| --- | --- | --- | --- | --- |
|  | Female |  | Male |  |
|  | Mean | Standard deviation | Mean | Standard deviation |
| Pre-implementation | 68 | 14 | 60 | 20 |
| Year 1 | 68 | 14 | 60 | 20 |
| Year 2 | 69 | 13 | 61 | 20 |
| Post- implementation | 69 | 14 | 61 | 20 |

<sup>a</sup>CPERS = Canadian Patient Experience Response Survey

**Table 2.** Ontario CPES-IC Response Rates - Overall and Measures (Discharge Communication & Planning and Discharge Management) by Fiscal Year

| Year | Number of responses | Response Rate |  |  |  |
| --- | --- | --- | --- | --- | --- |
|  |  | Number of Ontario hospitals<sup>a</sup> | Overall | Discharge communication & planning<sup>b</sup> | Discharge management<sup>c</sup> |
| 2016-2017 | 130,721 | 61 | 35.7 | 33.7 | 34.6 |
| 2017-2018 | 135,900 | 68 | 36.2 | 33.6 | 35.1 |
| 2018-2019 | 126,832 | 84 | 34.9 | 32.2 | 33.8 |
| 2019-2020 | 118,835 | 96 | 30.1 | 27.9 | 29.1 |

<sup>a</sup> n = Number of Ontario hospitals that submitted data to CPERS;
<sup>b</sup> The measure Discharge Planning consists of CPES-IC questions 19 (help needed after leaving hospital) and 20 (information in writing about symptoms to look out for);
°The measure Discharge Management consists of CPES-IC questions 37 (understanding about medications) and 38 (what to do if worried after leaving hospital) along with an additional question not included in our survey, question 39 (better understanding of condition post discharge).

Individuals discharged from hospitals with no PODS implementation had lower pre-implementation patient experience scores for all questions when compared to individuals discharged from hospitals with PODS implementation (Fig 1). The odds of reporting a positive patient experience in the post implementation period was statistically higher for two of four questions when compared to non-implementing hospitals (Table 3). Specifically, the odds of a positive response for having discussions with hospital staff on help needed and receiving information in writing about what symptoms to look for after leaving hospital was higher among hospitals with full implementation (cohort 1) when compared to those with no implementation (cohort 3). The odds of receiving information in writing about what symptoms to look for was also higher among partially implementing hospitals (cohort 2) when compared to those with no implementation (cohort 3). However, there was a statistically significant linear time trend difference between hospitals with full implementation (cohort 1) versus no implementation (cohort 3) for receiving information in writing about what symptoms to look for after leaving hospital and between hospitals with some implementation (cohort 2) versus no implementation (cohort 3) for having discussions with hospital staff on help needed (Table 3). Visual representations of the full model adjusting for time and clustering of sites with variability in the speed of each hospitals’ individual response are displayed in S1 Fig.

**Fig 1.**
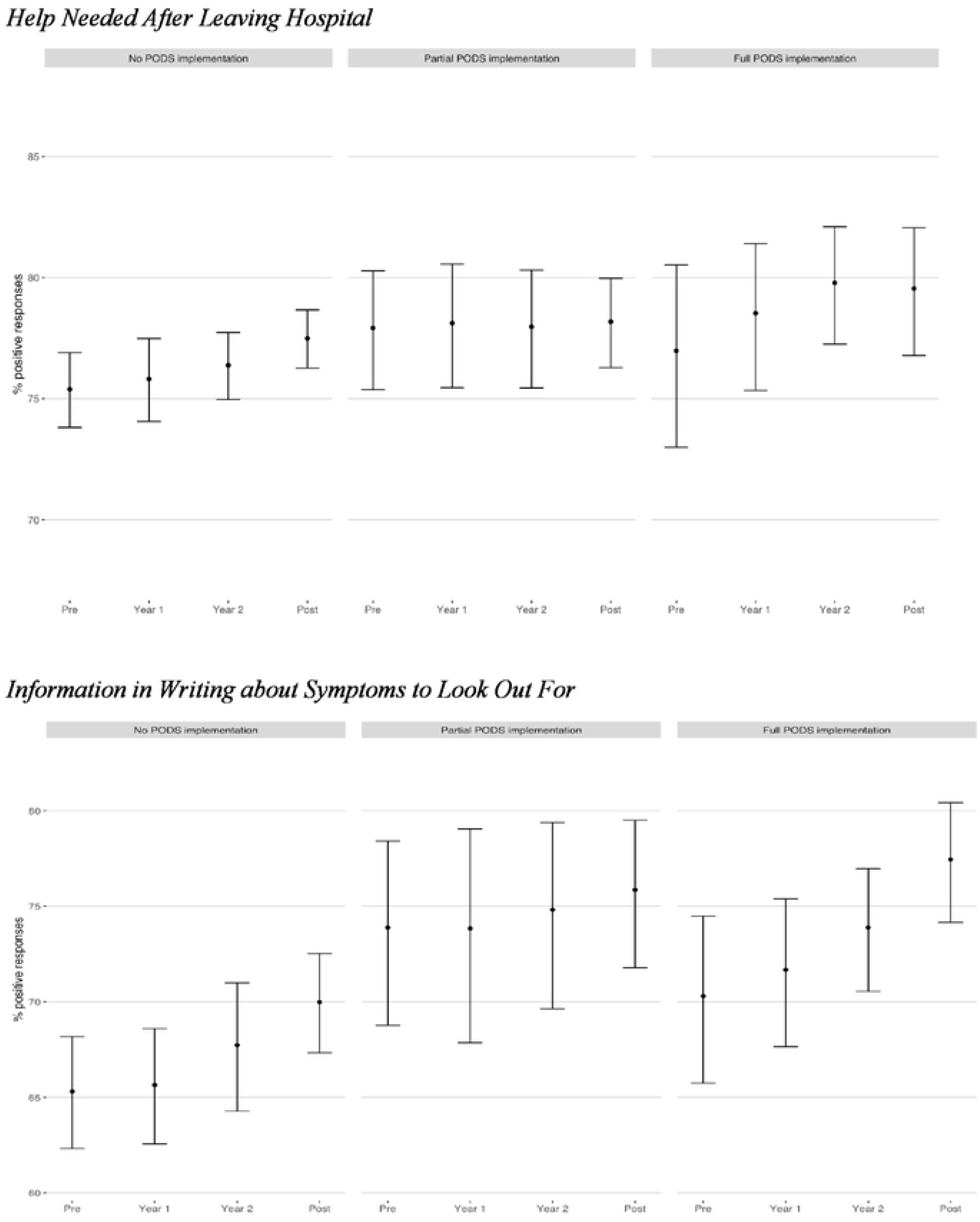
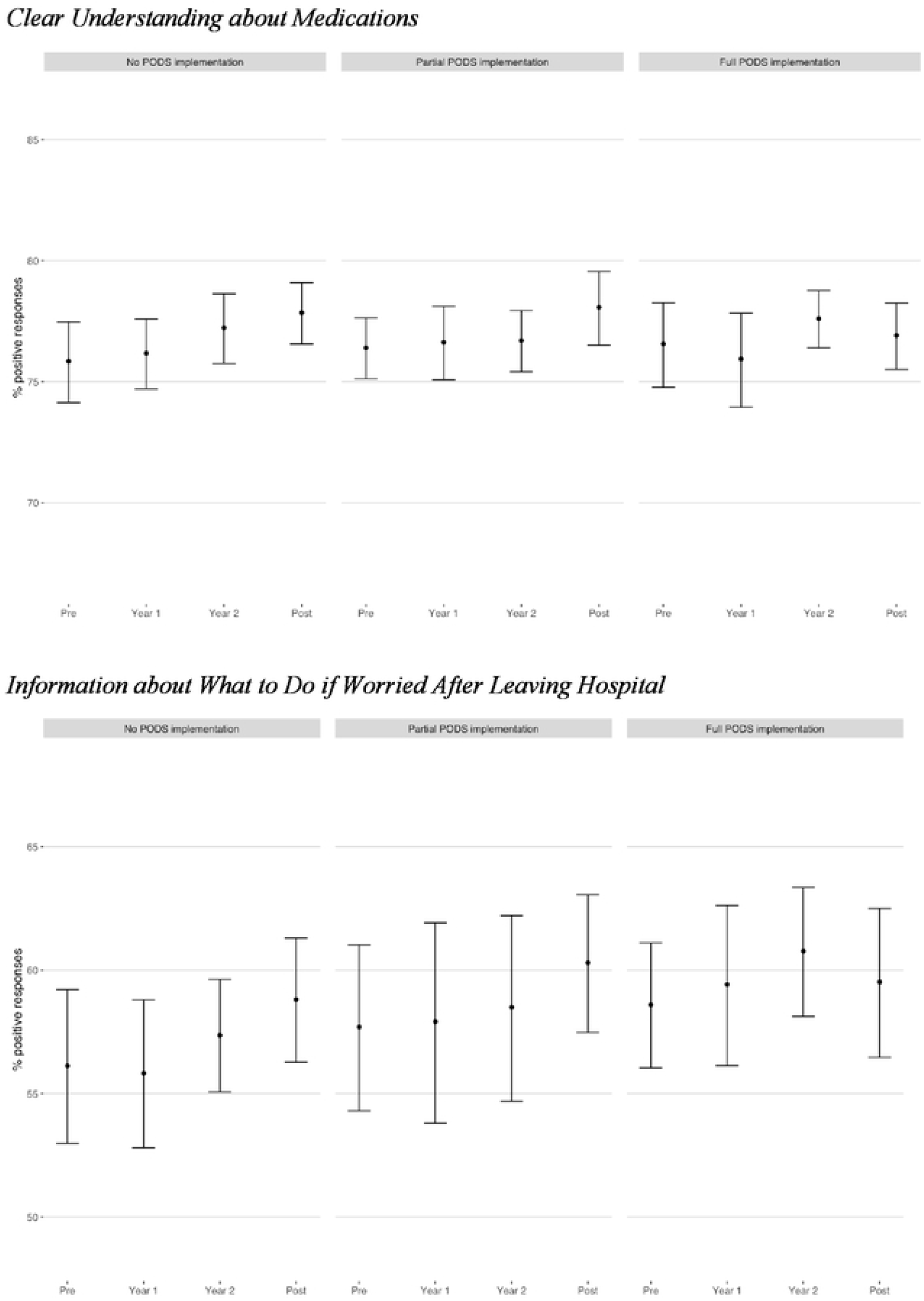
Percentages of positive patient experience by cohort over time, accounting for site level clustering, using GEE.

**Table 3.** Odds of a positive patient experience following discharge from acute care hospital one year following PODS implementation when compared to hospitals with no intervention

|  | **Full PODS**<br>**implementation** | **Partial PODS**<br>**implementation** |
| --- | --- | --- |

| Question | Odds ratio<br>(95% CI) | p-value | Odds Ratio<br>(95% CI) | p-value |
| --- | --- | --- | --- | --- |
| Help needed after leaving hospital | 1.18 (1.02-1.37) | 0.025 | 1.05 (0.92-1.20) | 0.488 |
| Information in writing about symptoms to look out for | 1.44 (1.17-1.78) | <0.001 | 1.35 (1.04-1.76) | 0.023 |
| Clear understanding about medications | 0.96 (0.86-1.08) | 0.517 | 0.99 (0.89-1.10) | 0.858 |
| Information about what to do if worried after leaving hospital | 1.08 (0.90-1.29) | 0.414 | 1.06 (0.91-1.22) | 0.473 |
| **Linear Time Trend (slope)** |  |  |  |  |
| Help needed after leaving hospital | 1.02 (0.98-1.06) | 0.440 | 0.97 (0.94-0.99) | 0.021 |
| Information in writing about symptoms to look out for | 1.05 (1.01-1.09) | 0.027 | 0.96 (0.91-1.02) | 0.209 |
| Clear understanding about medications | 0.98 (0.93-1.02) | 0.308 | 0.99 (0.95-1.02) | 0.507 |
| Information about what to do if worried after leaving hospital | 0.98 (0.92-1.04) | 0.480 | 0.99 (0.94-1.05) | 0.817 |

Note: There was a significant association between time and the outcome for cohort 3 for each outcome for the reference group (cohort 3) for all questions.

## Discussion

Our study found the delivery of a discharge instruction tool was associated with an improvement in patient experience for hospitals who implemented PODS, particularly for discharge planning. These are promising results given the recent attention both patients and Ontario Health, the province’s integrated health system planning and oversight agency, has given to improving quality standards in care transitions [16-18]. PODS however was not associated with an improvement on discharge management, particularly for understanding medications or what to do if worried after discharge, though linear time trends demonstrate active efforts in the province may be having a positive impact beyond the effect demonstrated by our tool. Our study highlights both where current care transition efforts are having the greatest impact and where gaps may still remain to improve patient experience.

Improving patient experience during care transitions from hospital to home has gained much attention over the last 10 years [4-9]. However, Canada has only focused on capturing patient experience recently [10,14,17-20]. Our study provides a deeper dive into areas for system improvement at a time when care coordination and communication practices were likely further hindered due to the COVID-19 pandemic [21,22]. Previous studies have demonstrated the role between high quality care transitions and post-discharge outcomes [1,3,9,12,23]. Interestingly, our study demonstrates higher patient experience scores than was reported in the only other cross-sectional study of patient experience across multiple Canadian provinces [20]. While some differences may be due to our focus on discharge management rather than patient satisfaction which was included in this study, our results likely represent the increasing attention on care transition quality and hospital-specific initiatives that are underway in Ontario [16]. The positive linear time trend reported in our non-implementing cohort is a reflection of these efforts. Our study strengthens prior work by providing repetitive cross-sectional measurement of patient experience over time allowing the identification of persistent gaps in patient experience across the largest Canadian province. Our study found a poor receipt of information on what to do if problems arise following hospitalization with just under 60% reporting positive scores. This finding may identify persistent gaps in this particular area but may also be a reflection of challenges in care coordination and health systems’ access patients’ and families’ have voiced they face once home [17].

While many interventions aim to help address poor post-discharge outcomes stemming from poor communication such as self-care behaviors or emergency department visits and readmissions [1-3], few have evaluated the impact on patient experience [24-27]. Our paper is one of the first to use Canadian patient experience measures to evaluate its association with the widespread implementation of a novel discharge communication tool. Our study is comparable to US studies which have studied the impact of care interventions on patient experience using identical or similar questions [26,27]. Patients randomized to receive a tailored discharge care plan along with one-on-one discussions with a health care provider on symptom recognition, medication reconciliation, and strategies for navigating the health system along with care coordination when needed were not found to have improved patient experience [26]. It is also possible that the impact of these care interventions lays more in the fidelity of the tool however –such as a higher engagement of caregivers or use of teach-back [13]. This may help explain why patients discharged from hospitals implementing PODS had a higher odds of having discussed the help they would need once leaving hospital, given patients may rely on caregivers for tasks beyond what public home care provides.

PODS was not found to improve patient experience measures related to discharge management. First, PODS was not found to improve the odds of reporting understanding of medications. In order to allow PODS to be usable across a wide range of patient populations and care systems, medication instructions were not standardized. Moreover, other province-wide care models centered on medication instructions were underway at the time of PODS implementation across Canada. Second, PODS was not found to improve understanding of what to do if problems arise after discharge from hospital. As a written form used prior to discharge to document instructions, PODS may do a better job of highlighting signs and symptoms patients and their families should watch out after leaving hospital, rather than where to seek care when complications arise. Prior studies have shown that seeking care when complications arise are influenced by system issues such as access to and relationships to primary care or specialist follow-ups, access issues which were unmeasured in our study [28,29]. Lastly, as PODS implementation did not include post-discharge reinforcements, care coordination or follow-up, this may help explain why questions centered on discharge management were not associated with implementation of our tool, unlike other care interventions [1].

Our study has several limitations. First, our study used hospital aggregate data, not individual patient data, and we cannot make causal inferences on the individual impact of receiving PODS on patient experience. While it is possible that individuals left a fully or partially implementing hospital with no PODS, this would make the likelihood of seeing an association less likely – and may have contributed in the partially implementing cohort. Second, the response rate for the survey is low across participating hospitals, with some hospitals having lower responses for certain time periods, and our results may not be representative of patient experience at all Ontario hospitals. This may be offset however by the wide representation of responses across both medical and surgical units and the similar low response rate in all cohorts which is consistent with response rates reported for this nationally used survey [10,20]. Future patient-level studies would benefit from a nonresponse adjustment being applied. Moreover, the likelihood that hospitals with the most interest in improving their patient experience were involved is high and may help explain why our study reported overall <u>higher</u> patient experience scores in all cohorts than reported previously [20]. Third, as the entire CPES-IC survey was not used in our study, it is possible we did not capture all aspects of care transition quality, and further research would be strengthened by the inclusion of patient-reported outcome measures. However, at minimum, we feel the chosen questions do reflect content areas in PODS and gaps in communication which have been identified in need of improvement [11,16,19].

Our study demonstrated an improvement to patient experience measures that center on discharge planning among individuals discharged home from Ontario medical and surgical units who implemented PODS. Our study highlights that while PODS is a promising discharge instruction tool, further refinement may be necessary in particular in areas which center on discharge management. Further research would benefit by including patient experience measures when evaluating new models of care or care interventions.

## Data Availability

Data cannot be shared publicly because they belong to many third parties. The data underlying the results presented in the study were made available to us by the Ontario Hospital Association in aggregate format. Data were given to us under the agreement that the raw data will not be shared. Others can access each dataset used in this study by approaching the respective institutions with letters from a recognized local ethics review committee and administrative authorities. We confirm that authors did not have any special access privileges to each dataset that others would not have.

## Acknowledgments

We would like to acknowledge Emily Myers, Program Lead of Patient Experience at the Ontario Hospital Association in providing valuable insights and data surrounding the CPES-IC Survey. We would also like to acknowledge our patient and caregiver advisory committee members: Audrey Chaput, Anna Foat, Gwen Cole, Trevor Manson and John Rae who collaborated in this work as outlined in the methods.

## Supporting information

S1 Fig. GEE model fits with site level data

## References

1. Jack BW, Chetty VK, Anthony D, Greenwald JL, Sanchez GM, Johnson AE, et al. A reengineered hospital discharge program to decrease rehospitalization: a randomized trial. Ann Intern Med. 2009 Feb;150(3):178–87. doi: 10.7326/0003-4819-150-3-200902030-00007

2. Williams MV, Li J, Hansen LO, Forth V, Budnitz T, Greenwald JL, et al. Project BOOST implementation: lessons learned. South Med J. 2014 Jul;107(7):455–65. doi: 10.14423/SMJ.0000000000000140

3. Coleman EA, Parry C, Chalmers S, Min SJ. The care transitions intervention: results of a randomized controlled trial. Arch Intern Med. 2006 Sep;166(17):1822–8. doi: 10.1001/archinte.166.17.1822

4. HCAHPS Hospital Consumer Assessment of Healthcare Providers and Systems. CAHPS hospital survey [Internet]. Baltimore, MD: Centers for Medicare & Medicaid Services; 2021 [updated 2021 Sep 30; cited 2021 Oct 1]. Available from: https://www.hcahpsonline.org

5. Graham MM, Norris CM. A new way to listen to patients: heeding patient reported experiences to improve quality of care. Can J Cardiol. 2019 Oct;35(10):1265–6. doi: 10.1016/j.cjca.2019.05.026

6. Aoki T, Yamamoto Y, Nakata T. Translation, adaptation and validation of the Hospital Consumer Assessment of Healthcare Providers and Systems (HCAHPS) for use in Japan: a multicentre cross-sectional study. BMJ Open. 2020 Nov 19;10(11):e040240. doi: 10.1136/bmjopen-2020-040240

7. Judan-Ruiz EA, Mina RJL, Macindo JRB. Psychometric properties of the Filipino version of Hospital Consumer Assessment of Healthcare Providers and Systems (HCAHPS): a cross-cultural validation study. J Patient Exp. 2020 Apr 2;7(6):1526–34. doi: 10.1177/2374373520912083

8. Zun AB, Ibrahim MI, Mokhtar AM, Halim AS, Wan Mansor WNA. Translation, cross-cultural adaptation, and validation of the Hospital Consumer Assessment of Healthcare Providers and Systems (HCAHPS) into the Malay language. Int J Environ Res Public Health. 2019 Jun 10;16(11):2054. doi: 10.3390/ijerph16112054

9. Coleman EA, Mahoney E, Parry C. Assessing the quality of preparation for posthospital care from the patient’s perspective: the care transitions measure. Med Care. 2005 Mar;43(3):246–55. doi: 10.1097/00005650-200503000-00007

10. Hadibhai S, Lacroix J, Leeb K. Developing the first pan-Canadian acute care patient experience survey. Patient Exp J. 2018;5(3):25–33. doi:10.35680/2372-0247.1227

11. Hahn-Goldberg S, Okrainec K, Huynh T, Zahr N, Abrams H. Co-creating patient-oriented discharge instructions with patients, caregivers, and healthcare providers. J Hosp Med. 2015 Dec;10(12):804–7. doi: 10.1002/jhm.2444

12. Oh EG, Lee HJ, Yang YL, Kim YM. Effectiveness of discharge education with the teach-back method on 30-day readmission: a systematic review. J Patient Saf. 2021 Jun;17(4):305–10. doi: 10.1097/PTS.0000000000000596

13. Hahn-Goldberg S, Huynh T, Chaput A, Krahn M, Rac V, Tomlinson G, et al. Implementation, spread and impact of the Patient Oriented Discharge Summary (PODS) across Ontario hospitals: a mixed methods evaluation. BMC Health Serv Res. 2021 Apr 17;21(1):361. doi: 10.1186/s12913-021-06374-8

14. Canadian Institute for Health Information. Patient experience in Canadian hospitals – methodology notes [Internet]. Ottawa, ON: CIHI; 2019 [cited 2021 Mar 15]. Available from: https://secure.cihi.ca/free_products/cpes-patient-experience-methodology-notes-april2019-en-web.pdf

15. von Elm E, Altman DG, Egger M, Pocock SJ, Gotzsche PC, Vandenbroucke JP. The Strengthening the Reporting of Observational Studies in Epidemiology (STROBE) Statement: guidelines for reporting observational studies. J Clin Epidemiol. 2008 Apr;61(4):344–9. doi: 10.1016/j.ijsu.2014.07.013

16. Health Quality Ontario. Evidence to improve care – quality standards [Internet]. Toronto, ON: HQO; date unknown [updated 2021; cited 2021 Jul 29]. Available from: https://www.hqontario.ca/Evidence-to-Improve-Care/Quality-Standards/View-all-Quality-Standards/Transitions-between-hospital-and-home/About

17. Kiran T, Wells D, Okrainec K, Kennedy C, Devotta K, Mabaya G, et al. Patient and caregiver experience in the transition from hospital to home - brainstorming results from group concept mapping: a patient-oriented study. CMAJ Open. 2020 Mar 2;8(1):e121–e133. doi: 10.9778/cmajo.20190009

18. Kemp KA, Santana MJ, Southern DA, McCormack B, Quan H. Association of inpatient hospital experience with patient safety indicators: a cross-sectional, Canadian study. BMJ Open. 2016 Jul 1;6(7):e011242. doi: 10.1136/bmjopen-2016-011242

19. Kiran T, Wells D, Okrainec K, Kennedy C, Devotta K, Mabaya G, et al. Patient and caregiver priorities in the transition from hospital to home: results from province-wide group concept mapping. BMJ Qual Saf. 2020 May;29(5):390–400. doi: 10.1136/bmjqs-2019-009993

20. Davidson MJ, Lacroix J, McMartin S, Kudhail R, Hart K, Diestelkamp N. Patient experiences in Canadian hospitals. Healthc Q. 2019 Oct;22(3):12–4. doi: 10.12927/hcq.2019.26024

21. Key T, Kulkarni A, Kandhari V, Jawad Z, Hughes A, Mohanty K. The patient experience of inpatient care during the COVID-19 pandemic: exploring patient perceptions, communication, and quality of care at a university teaching hospital in the United Kingdom. J Patient Exp. 2021 Mar 3;8:2374373521997742. doi: 10.1177/2374373521997742

22. Verma AA, Hora T, Jung HY, Fralick M, Malecki SL, Lapointe-Shaw L, et al. Characteristics and outcomes of hospital admissions for COVID-19 and influenza in the Toronto area. CMAJ. 2021 Mar 22;193(12):e410–8. doi: 10.1503/cmaj.202795

23. Hahn-Goldberg S, Jeffs L, Troup A, Kubba R, Okrainec K. “We are doing it together”; the integral role of caregivers in a patients’ transition home from the medicine unit. PLoS ONE. 2018 May 24;13(5):e0197831. doi: 10.1371/journal.pone.0197831

24. Backman C, Chartrand J, Crick M, Burry RD, Dingwall O, Shea B. Effectiveness of person-and family-centred care transition interventions on patient-oriented outcomes: A systematic review. Nurs Open. 2021 Mar;8(2):721–54. doi: 10.1002/nop2.677

25. Okrainec K, Lau D, Abrams HB, Hahn-Goldberg S, Brahmbhatt R, Huynh T, et al. Impact of patient-centered discharge tools: A systematic review. J Hosp Med. 2017 Feb;12(2):110–7. doi: 10.12788/jhm.2692

26. Chan B, Goldman LE, Sarkar U, Schneidermann M, Kessell E, Guzman D, et al. The effect of a care transition intervention on the patient experience of older multi-lingual adults in the safety net: results of a randomized controlled trial. J Gen Intern Med. 2015 Dec;30(12):1788–94. doi: 10.1007/s11606-015-3362-y

27. Kangovi S, Mitra N, Grande D, White ML, McCollom S, Sellman J, et al. Patient-centered community health worker intervention to improve posthospital outcomes: a randomized clinical trial. JAMA Intern Med. 2014;174(4):535–43. doi: 10.1001/jamainternmed.2013.14327

28. Lam K, Abrams HB, Matelski J, Okrainec K. Factors associated with attendance at primary care appointments after discharge from hospital: a retrospective cohort study. CMAJ Open. 2018 Dec 3;6(4):e587–e593. doi: 10.9778/cmajo.20180069

29. Griffiths S, Stephen G, Kiran T, Okrainec K. “She knows me best”: a qualitative study of patient and caregiver views on the role of the primary care physician follow-up post-hospital discharge in individuals admitted with chronic obstructive pulmonary disease or congestive heart failure. BMC Fam Pract. 2021 Sep 7;22(1):176. doi: 10.1186/s12875-021-01524-7

